# Joining the Conversation with a Dedicated Medical Education Corpus

**DOI:** 10.1101/2024.12.17.24319205

**Authors:** Gregory M. Ow, Geoffrey V. Stetson, Joseph A. Costello, Anthony R. Artino, Lauren A. Maggio

## Abstract

**PROBLEM:** Medical education scholars struggle to join ongoing conversations in their field due to the lack of a dedicated medical education corpus. Without such a corpus, scholars must either search too widely across thousands of irrelevant journals, or too narrowly by relying on PubMed’s Medical Subject Headings (MeSH). In our tests, MeSH missed 34% of medical education papers.

**APPROACH:** We developed the MEC (Medical Education Corpus), the first dedicated collection of medical education papers, through a three-step process. First, using the Core-Periphery model, we created the MEJ (Medical Education Journals), a collection of three groups of journals based on participation and influence in medical education discourse: the MEJ-Core (formerly the MEJ-24, 24 journals), MEJ-Adjacent (127 journals), and MEJ-Peripheral (a theoretical group). Second, we developed and evaluated a machine learning model, the MEC Classifier, trained on 4,032 manually labeled papers to identify medical education content. Finally, we applied the MEC Classifier to extract medical education papers from the MEJ-Core and MEJ-Adjacent journals.

**OUTCOMES:** The MEC currently contains 119,137 medical education papers from the MEJ-Core (54,927 papers) and MEJ-Adjacent journals (64,210 papers). In our evaluation using 1,358 test papers, the MEC Classifier demonstrated significantly improved sensitivity compared to MeSH (90% vs 66%, *p* = 0.001), even while maintaining a similar positive predictive value (82% vs 81%).

**NEXT STEPS:** The MEC provides a focused, searchable corpus that enables medical education scholars to more easily join conversations in the field. Scholars can now rely on the MEC when reviewing literature to frame their work, and the MEC also creates opportunities for field-wide analyses and meta-research. The MEC is freely available in the Supplement and is regularly updated on MedEdMentor (mededmentor.org), where we will also incorporate community feedback to further improve and expand the corpus.

## Problem

When writing their manuscripts, medical education scholars are urged to join ongoing conversations in the field — to listen, then contribute something relevant and new.^1^ However, scholars struggle with the “listening” process, which involves reviewing the existing medical education literature to accurately frame their work.^2^ This struggle partially stems from the lack of a defined literature base. After all, if scholars cannot easily locate the field’s discourse, how can they contribute to the conversation? To address this issue, we introduce the MEC (Medical Education Corpus), a growing collection of 119,137 medical education papers extracted from 151 medical education journals. The MEC is searchable via the MedEdMentor website,^3^ and a snapshot of the corpus saved in December 2024 is downloadable in the Supplement.^4^

If medical education papers are contributions to a conversation, and journals exist to promote these conversations,^1^ then the MEC is an academic conference — a dedicated space that brings these conversations together. Without a corpus like the MEC, scholars face two competing problems. The first problem arises from searching too broadly. Scholars frequently search biomedical databases like PubMed, or general academic search engines like Google Scholar. But these resources are composed of medical education journals mixed in with thousands of unrelated journals. Like trying to listen to a quiet conversation in a bustling conference lobby, these searches require significant effort to separate the medical education discourse from the background noise. The second problem arises from searching too narrowly. To our knowledge, the only systematic method to identify medical education papers is PubMed’s Medical Subject Headings (MeSH) — a controlled vocabulary thesaurus used for indexing papers in the MEDLINE subset of PubMed.^5^ Yet in our tests, MeSH missed 34% of medical education papers, thereby inadvertently muffling a significant portion of the conversation.

The MEC addresses both of these problems by assembling medical education conversations into one metaphorical conference. At this conference, scholars can hear conversations with much less noise (the MEC only draws from medical education journals), and substantially fewer missed conversations (the MEC misses just 10% of medical education papers in our tests). As a result, the MEC enables scholars to more effectively join the conversations that matter to their scholarship.

## Approach

### Overview

We developed the MEC in three steps. First, we created a set of medical education journals to extract from. Second, we developed and evaluated a machine learning model to identify medical education papers. Finally, we used our model to extract the medical education papers from our journals, thus forming the MEC (Figure 1).

**Figure 1.**
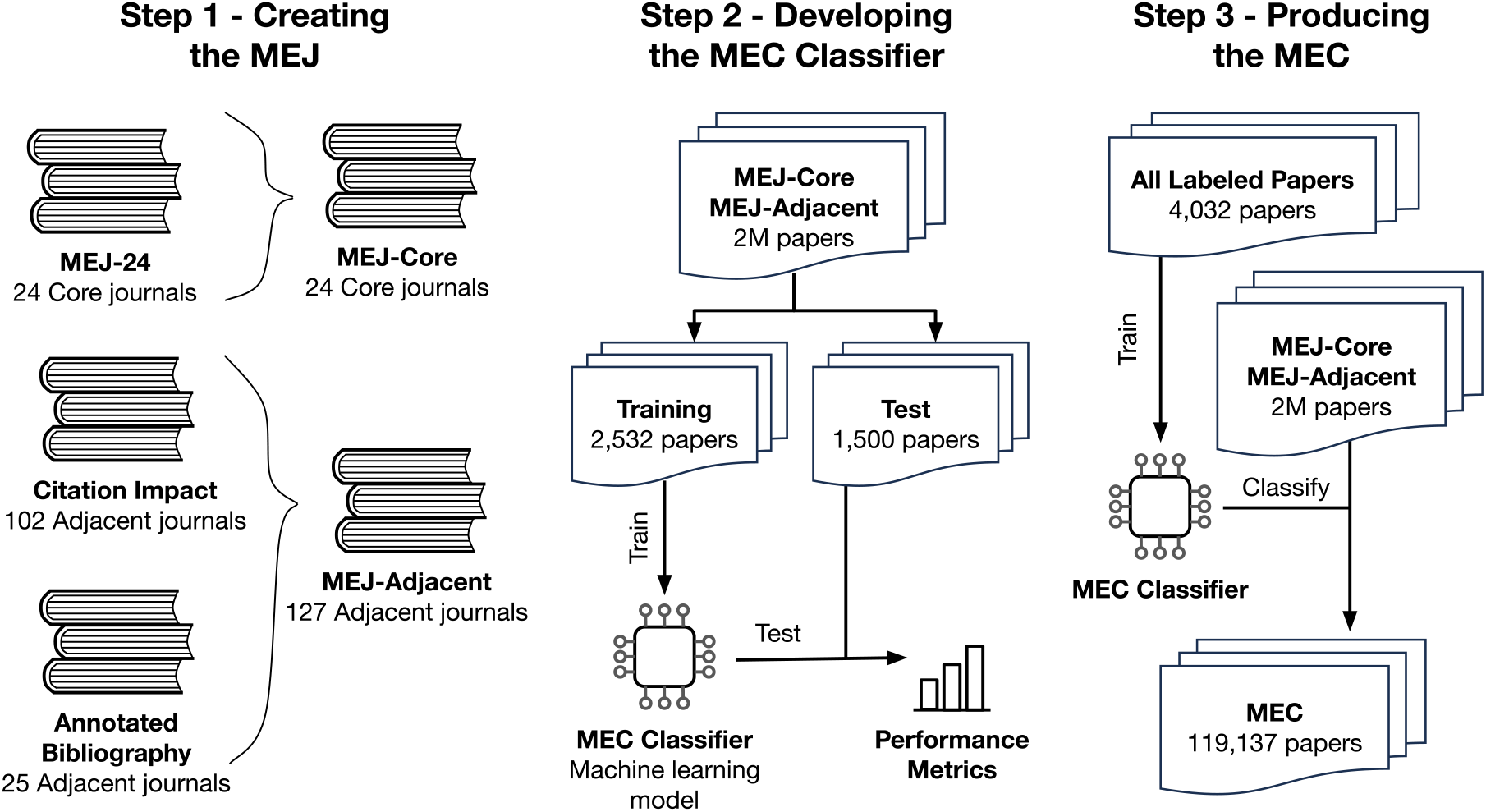
Development process of the Medical Education Corpus (MEC). The three-step process used to create the MEC: (1) Creating three groups of medical education journals, collectively named the MEJ (Medical Education Journals); (2) Developing the MEC Classifier by training and evaluating a machine learning model; and (3) Producing the final MEC by applying the MEC Classifier on the MEJ-Core and MEJ-Adjacent journals, resulting in the MEC.

### Step 1 - Creating a set of medical education journals to extract from

As of 2010, 4,208 journals had published at least one medical education paper.^5^ As a result, prior efforts to identify “medical education journals” have focused only on the journals *most* dedicated to medical education.^5,6^ However, since we designed our machine learning model (see Step 2) to extract medical education papers from even the *least* dedicated journals, we adopted a more expansive approach.

We begin by adapting the Core-Periphery model from network theory^7^ to organize and enumerate the most significant medical education journals. This model describes densely interconnected Core members loosely connected to Peripheral members. Applying this model, we created three groups of journals, collectively named the MEJ (Medical Education Journals). The MEJ consists of:

1. **MEJ-Core —** this group of journals consistently publishes medical education papers and forms a tight-knit citation network among themselves (e.g. *Academic Medicine, Medical Education*).
2. **MEJ-Adjacent** — this group of journals occasionally publishes medical education papers and are moderately cited from the Core (e.g. *New England Journal of Medicine*).
3. **MEJ-Peripheral** — this group of journals rarely publishes medical education papers and are only sparsely cited from the Core. The MEJ-Peripheral currently exists only as a theoretical group. Given the impracticality of identifying the thousands of journals that rarely produce medical education content, we did not enumerate this group for this study.

The MEJ-Core currently consists of the MEJ-24 (Medical Education Journals-24), a set of 24 central medical education journals assembled using bibliometrics.^6^ With input from several originators of the MEJ-24 (LM, JC, AA), we establish the MEJ-Core as a more-flexibly-named set that can expand beyond 24 journals as medical education evolves.

The MEJ-Adjacent consists of 127 journals (see Supplement) identified through two criteria, excluding journals already in the MEJ-Core:

1. **Citation Impact (102 journals) —** We utilized a prior analysis to identify journals cited 500+ times in MEJ-Core papers between 2000-2020.^8^ This threshold was chosen by author consensus to identify journals that regularly engaged with medical education. Notably, *Science* had 1,120 citations, but was excluded given its large size (∼250,000 papers) with no medical education papers found on PubMed since 2019.
2. **Expert Opinion (25 additional journals) —** To ensure we also leveraged expert opinion, we added the journals in the Annotated Bibliography, a resource published by AAMC’s Medical Education Scholarship Research and Evaluation Section.^9^ This resource lists journals that publish health professions education manuscripts.

Together, the MEJ-Core and MEJ-Adjacent represent the 151 journals from which we will extract medical education papers.

### Step 2 - Developing and evaluating a machine learning model to identify medical education papers

We developed the MEC Classifier, a machine learning model that identifies medical education papers based on their titles, abstracts, and journals. Machine learning models are computer algorithms that “learn” patterns from data.

To train the model, we first gathered data for the model to learn from. In March 2024, we retrieved all 2.2 million papers in the MEJ-Core and MEJ-Adjacent from Semantic Scholar, an openly-accessible index of academic papers. From these 2.2 million papers, we strategically selected 2,532 papers that would be the most informative examples for the model to learn from (i.e. active learning, see Supplement). Each paper received two independent labels from our author team — composed of experienced medical librarians (LM, JC), clinician-educators (GO, GS), and medical education scholars (AA, LM). Papers were labeled as medical education if they met any of the following criteria: (1) focused on undergraduate, graduate, or continuing medical education; (2) involved premedical students, medical students, interns, residents, or fellows; or (3) otherwise appreciably pertained to medical education (e.g. surgical simulation). Full texts were accessed as needed, and discordant labels were resolved by obtaining a tiebreaking label.

To evaluate the MEC Classifier’s performance, we created an independent set of test papers. In October 2024, we randomly sampled an additional 1,500 papers from the MEJ-Core and MEJ-Adjacent for testing — these papers were not part of the training data. The test papers were labeled as above, and our labels served as the “answer key” for the test. We then trained the MEC Classifier on the initial 2,532 training papers, and then used it to classify the 1,500 test papers. To assess the model’s performance, we compared its classifications against our answer key (results in Outcomes).

To contextualize the MEC Classifier’s performance, we compared it against PubMed’s MeSH. For a fair comparison, we used only the subset of our test papers that were available on PubMed (1,358/1,500 test papers, 91%). We evaluated two MeSH-based approaches: the “Education, Medical” MeSH term alone,^5^ and the broader set of all medical-education-related terms (“Education, Medical” OR “Schools, Medical” OR “Students, Medical” OR “Faculty, Medical”).

### Step 3 - Producing the MEC

We retrained the MEC Classifier on all 4,032 labeled papers (2,532 training papers and 1,500 test papers) to maximize its effectiveness. In December 2024, we used this final version to extract all medical education papers from the MEJ-Core and MEJ-Adjacent, producing the MEC.

All labeled papers, the code used to train the MEC Classifier, and additional technical details are provided in the Supplement. This information is sufficient to recreate the MEC Classifier, which can be used to create the MEC.

## Outcomes

### The MEJ consists of three groups of medical education journals

The MEJ is a collection of three groups of journals: the MEJ-Core (24 journals), MEJ-Adjacent (127 journals), and MEJ-Peripheral. Analysis of citations from MEJ-Core papers revealed that 40.8% of their citations were to other papers in MEJ-Core journals, and 31.0% were to papers in MEJ-Adjacent journals.^8^ This validates our choice to build the initial MEC from these journals, as they account for the majority of the field’s scholarly engagement.

### The MEC is extracted from the MEJ-Core and MEJ-Adjacent

From all 2,259,771 MEJ-Core and MEJ-Adjacent papers as of December 2024, our final model identified 119,137 as medical education papers (5.3%). These papers constitute the MEC.

There were 54,927 medical education papers from 64,572 MEJ-Core papers (85%), and 64,210 medical education papers from 2,195,199 MEJ-Adjacent papers (2.9%).

### The MEC Classifier effectively identifies medical education papers

The value of the MEC derives from how well the MEC Classifier can identify medical education papers.

In our 1,500 test papers from the MEJ-Core and MEJ-Adjacent, the MEC Classifier was highly effective. It correctly identified 86% (68/79) of medical education papers (i.e. 86% sensitivity). And it was relatively discerning: when it identified a paper as medical education, it was correct 81% (68/84) of the time (i.e. 81% positive predictive value, PPV).

In the subset of test papers available on PubMed (1,358 papers), the MEC Classifier was superior to MeSH terms (Figure 2). The MEC Classifier correctly captured 90% of medical education papers while MeSH managed just 66%, despite combining all medical-education-related terms (McNemar’s χ^2^ = 10.24, *p* = 0.001). Even while capturing more papers, the MEC Classifier was just as discerning as MeSH: both had a PPV around 80%.

**Figure 2.**
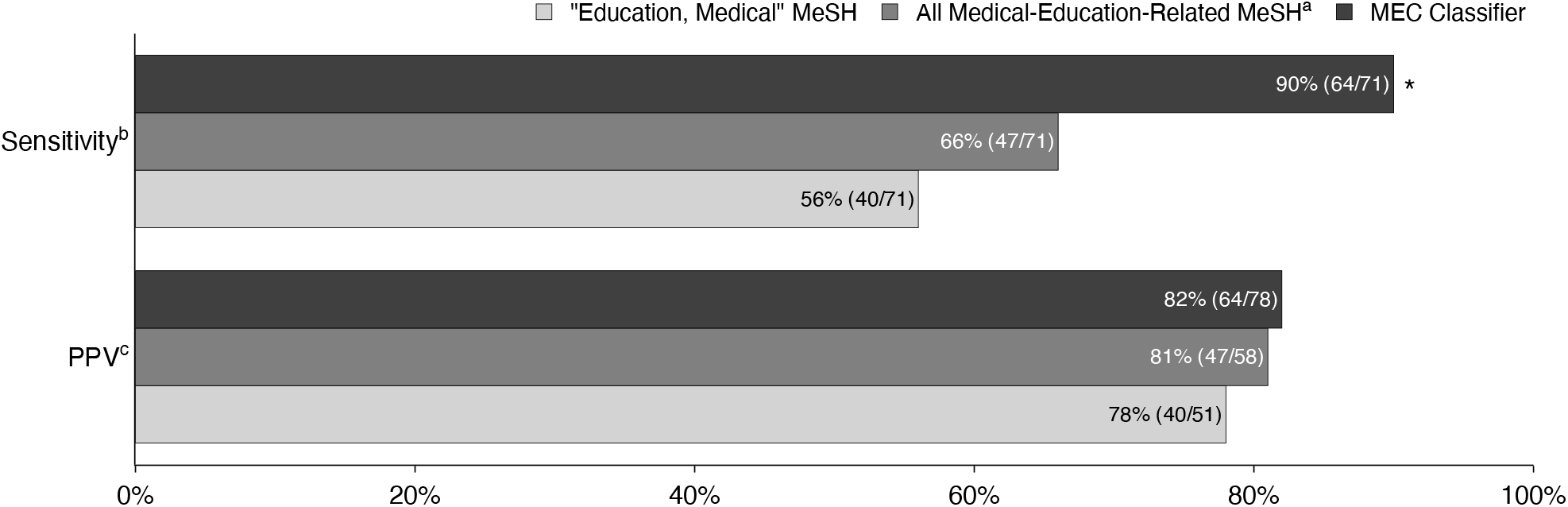
Performance metrics comparison between the MEC Classifier and MeSH. Comparison between three different methods for classifying medical education papers. Bar chart shows the sensitivity and positive predictive value for: the MEC Classifier (black bars), the combination of all medical-education-related MeSH terms (dark gray bars), the “Education, Medical” MeSH term alone (light gray bars).^a^ Analysis performed on 1,358 randomly sampled papers from the MEJ-Core and MEJ-Adjacent journals that were available on PubMed in October 2024. ^a^All medical-education-related MeSH terms are composed of ("Education, Medical" OR "Schools, Medical" OR "Students, Medical" OR "Faculty, Medical"). ^b^Sensitivity is the proportion of medical education papers correctly identified ^c^PPV (Positive Predictive Value) is the proportion of papers classified as medical education that were actually medical education. ^*^McNemar's X^2^ = 10.24, *p* = 0.001, when compared against all medical education-related MeSH.

### Compared to MeSH, the MEC Classifier includes more medical education papers from major journals

The journal-level impacts of these differences in sensitivity are substantial. The MEC Classifier captures a much larger fraction of major medical education journals like *MedEdPORTAL* and *Teaching and Learning in Medicine* (Figure 3) — in some cases capturing an additional one-third of a journal’s papers. For *Academic Medicine*, the MEC Classifier identified 73% of articles as medical education, similar to the 70% identified when all medical-education-related MeSH terms were used (see Supplement).

**Figure 3.**
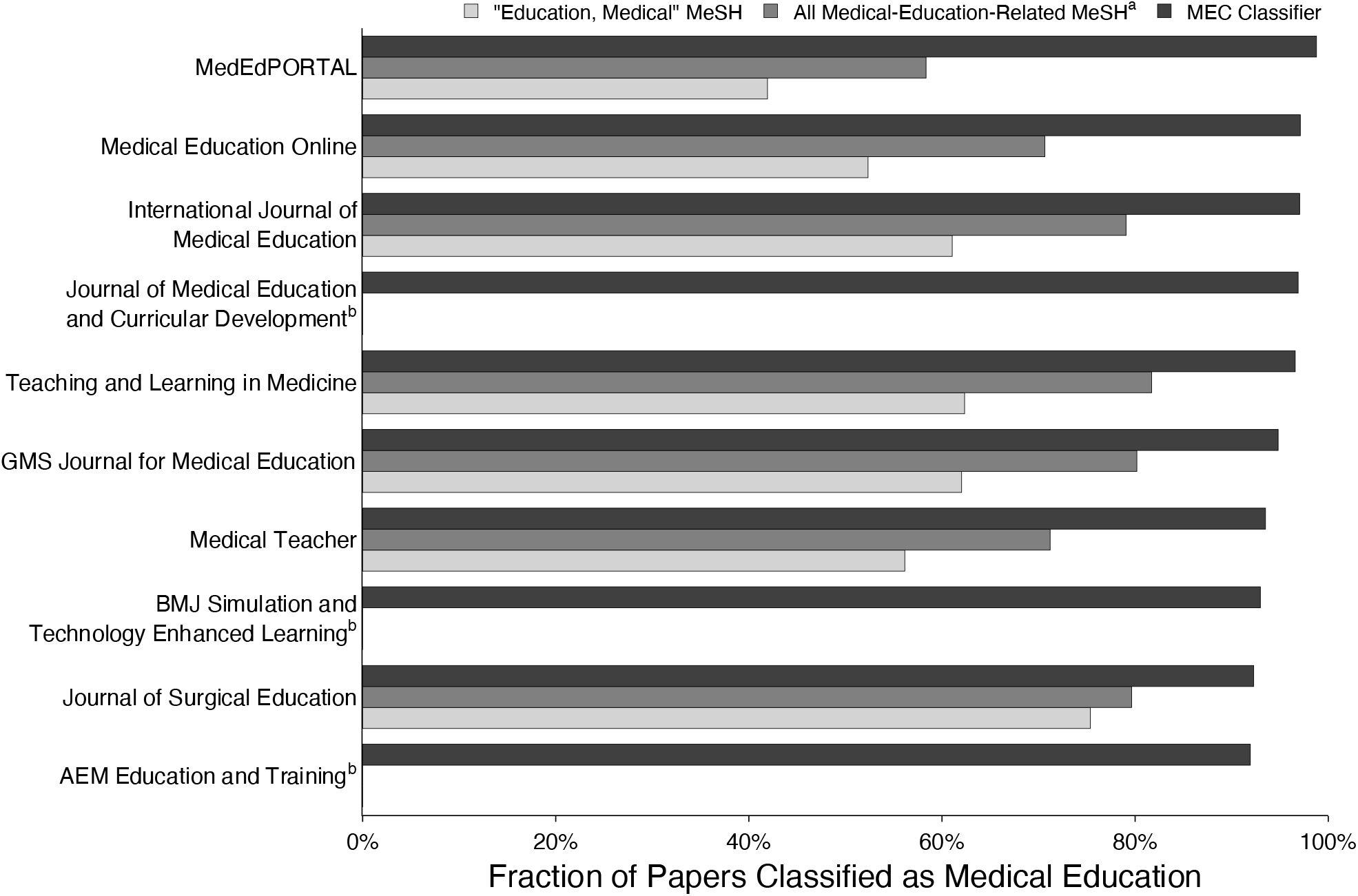
MEC Classifier versus MeSH by journal. Comparison across the 10 journals with the highest proportion of medical education content (as identified by the MEC Classifier). Bar chart showing the fraction of papers classified as medical education for: the MEC Classifier (black bars), the combination of all medical-education-related MeSH terms (dark gray bars), the “Education, Medical” MeSH term alone (light gray bars).^a^ Analysis performed on the 2.3 million papers published in MEJ-Core and MEJ-Adjacent journals as of December 2024. ^a^All medical-education-related MeSH terms are composed of ("Education, Medical" OR "Schools,
Medical" OR "Students, Medical" OR "Faculty, Medical"). ^b^*Journal of Medical Education and Curricular Development, BMJ Simulation and Technology Enhanced Learning, and AEM Education and Training* are not MEDLINE-indexed and so do not receive MeSH labels.

## Next Steps

The MEC assembles medical education papers into one searchable corpus. Scholars can more easily join conversations, and meta-researchers can analyze the field (e.g. study trends in theory usage). Like a well-organized conference, the MEC carefully selects its program, only drawing from the 151 journals currently in the MEJ. Furthermore, the MEC captures 90% of medical education papers from these journals compared to MeSH’s 66% — so more conversations are heard.

Practically, these qualities mean that searching the MEC alone will suffice for straightforward purposes like reviewing existing literature to frame scholarly work. However, for more comprehensive tasks, scholars still require advanced search strategies and librarian expertise. In particular, scholars conducting systematic reviews should *not* rely solely on the MEC. The MEC’s current exclusion of MEJ-Peripheral journals means it cannot provide the comprehensive literature coverage such reviews require.^10^

To address two limitations in the MEC’s development, we will implement community feedback processes through MedEdMentor. The first limitation arises from MEJ’s citation-based journal grouping, which favors established journals over newer or historically marginalized journals. To address this, community members will be able to suggest journals to add to the MEJ. The second limitation relates to potential biases in our team’s definition of what constitutes a “medical education” paper. To address this, we will enable qualified community members to provide paper labels, which will be used to periodically update the MEC Classifier.

The MEC lays the foundation for a more inclusive scholarly conversation in medical education. We believe the corpus will be especially valuable to scholars with limited access to experienced librarians and mentors. To promote accessibility, the MEJ and a snapshot of the MEC are available to download in the Supplement. Because the MEC is derived from published papers, it is a dynamic corpus that grows as new papers are published. Therefore it is regularly updated and searchable on MedEdMentor.

## Data Availability

Technical Supplement to the Medical Education Corpus - https://doi.org/10.5281/zenodo.14511550

## Acknowledgements

The authors would like to sincerely thank Alex Iyer for his thoughtful review and feedback on an early draft of this manuscript.

